# Illicit Cigarette Trade and Tax evasion in Zambia: Findings from the Tobacco Control Data Initiative 2023

**DOI:** 10.1101/2024.10.14.24315433

**Authors:** Cosmas Zyambo, Masauso Moses Phiri, Webby Mwamulela, Richard Zulu, Mbaita Maka, Aminata Camara, Sharon Ogolla, Seember Joy Ali, Retselisitsoe Pokothoane, Hana Ross, Fastone Matthew Goma, Noreen Dadirai Mdege

## Abstract

**Background:** Illicit cigarette trade has significant economic and public health implications. It leads to governments tax revenue losses due to the evasion of tobacco taxes, and often these cigarettes are cheaper ones therefore increasing cigarette consumption.

**Objective:** To estimate the Illicit cigarette trade and tax evasion in Zambia and establish its associated factors

**Methods and analysis:** A cross sectional survey was used to collect empty cigarette packs from the retailers and street/bins in 25 districts covering 10 Provinces of Zambia. We used a descriptive analysis to calculate the proportion of illicit cigarette packs and other specific criteria. Logistic regression was used to model the factors associated with the prevalence of the illicit cigarette market in Zambia.

**Results:** Of the 118, 344 empty cigarette packs collected (82.0% from the retailers and 18.0% the street/bins), Rothmans accounted for 40.7% and Stuyvesant 13.1%, both manufactured by British American Tobacco. 14,428 (12.2%) were deemed illicit. Out of the total packs, 1792 (1.5%) did not have a textual health warning, 343 (0.3%) packs did not have a textual health warning in english, 1490 (1.3%) had duty-free stamps even though they were purchased from retail outlets that were not duty-free shops and, 11,939 (10.1%) did not have a ZRA stamp. Factors associated with reduced odds of illicit cigarettes sales were non-boarder [AOR 0.17 (CI; 0.13 – 0.23)] and local manufactured AOR 0.44 (CI; 0.37 – 0.53).

**Conclusions:** Our study demonstrated that 12.2% of the cigarettes sold on the Zambian market is illicit, with 10.1% evading tax. We found that cigarettes from Lusaka province, urban regions, border towns, and those that are imported had higher odds of being illicit. This finding underscores the fact that Zambia should ratify and implement the WHO Protocol on Illicit Tobacco Trade (ITP) to counter the supply of illicit cigarettes.

## Introduction

Tobacco use is a major risk factor to six of the eight leading causes of death in the world ^1,2^. Zambia has a smoking prevalence of about 12.3%, with data demonstrating an increase, particularly in men from 15% in 2000-2002 to 19% in 2018^3–5^. It is estimated that 7,142 people lose their lives due to tobacco-related diseases annually in Zambia, mostly from heart diseases, cancers, and other non-communicable diseases (NCDs)^6,7^. Tobacco use also costs the government of Zambia 2.8 billion Zambian Kwacha (ZMW), which is 1.2% of its GDP, annually^6^. This includes 154 million ZMW in healthcare expenditure and 2.7 billion ZMW in lost productivity from premature mortality, disability and workplace smoking breaks^6^.

The World Health Organization Framework Convention on Tobacco Control (WHO FCTC) provides a number of evidence-based policy interventions that can be implemented to address the tobacco epidemic and its consequences. Unfortunately, illicit trade of cigarettes undermines tobacco control policies by increasing access to cigarettes, often cheaper ones, and therefore increasing cigarette consumption^8–11^. It also reduces government tax revenues. Article 15 of WHO-FCTC requires Parties to the Convention to implement effective measures against all forms of illicit trade in tobacco products including smuggling, illicit manufacturing, and counterfeiting. The WHO-FCTC adopted the Protocol to Eliminate Illicit Trade in Tobacco Products in 2012 to combat illicit trade in tobacco products by securing the supply chain with measures such as track and trace systems^6,12^. It also covers licensing, due diligence, and issues related to internet- and telecommunication-based sales, tobacco product transactions in free zones and international transit, as well as duty free sales.

Although Zambia became a Party to the WHO FCTC in 2008^6^, it has not fully implemented article 15 on illicit trade tobacco products and is currently not a signatory to the Protocol to Eliminate Illicit Trade in Tobacco Products. Zambia, like many developing countries, does not have estimates of the size of the illicit trade market of cigarettes that are independent of the tobacco industry. Tobacco Institute of Southern Africa in Zambia estimates that there were more than 400 million cigarettes that entered the market through smuggling, counterfeiting or tax evasion annually and this accounted for 30% of the cigarette market^13,14^. Unfortunately, the tobacco industry is known for exaggerating the magnitude of the illicit cigarette with the sole purpose of preventing the governments from raising the taxes and prices of tobacco products. There is therefore a need for context-specific studies on the extent and nature of illicit cigarette trade in Zambia that are independent of the tobacco industry. Such studies are vital in informing the development and implementation of context-specific tobacco control policies and interventions to reduce access to and consumption of cigarettes. We therefore aimed to estimate the proportion of cigarettes consumed that are illicit, the extent of cigarette tax evasion, and the associated factors in Zambia.

## Methodology

### Data Source

This was an observational cross-sectional study based on empty cigarette packs collected between November and December 2022. By law, the sale of single cigarette sticks is not prohibited in Zambia and is highly prevalent. We therefore approached retailers and street vendors in the study areas who sell cigarettes and requested them to keep any empty cigarette packs from their sales for one day (i.e., pack collection day). The research team collected the empty packs at the close of business on pack collection day. Additionally, we also collected empty cigarette packs that were discarded on the streets, near garbage bins, and waste dumps.

### Sampling

Zambia is divided into 10 provinces and 116 districts. Zambia also has large international land borders with eight neighboring countries (Figure 1). From each of the ten provinces in Zambia, we stratified the districts into border and non-border districts. A border district was defined as a district with a formal border linking Zambia to one of its neighboring countries. We randomly selected 20 districts to be included in the study. We also purposely added the following busiest commercial border districts that had not been randomly selected to our sample: Chililabombwe and Mufulira districts which border with the Democratic Republic of Congo, and Chirundu district bordering with Zimbabwe. We also purposely included two districts with big cities, Lusaka and Chipata, to the sample. Thus, a total of 25 districts, which included 9 border districts and 16 non-border districts covering all 10 Provinces, were selected for the study. Among these 25 districts were five urban districts, i.e., those falling under municipalities, and 20 rural districts, i.e., those outside municipalities. Within each district, we selected the central business district (CBD) and identified the main market within the CBD as the starting point for data collection.

**Figure 1:**
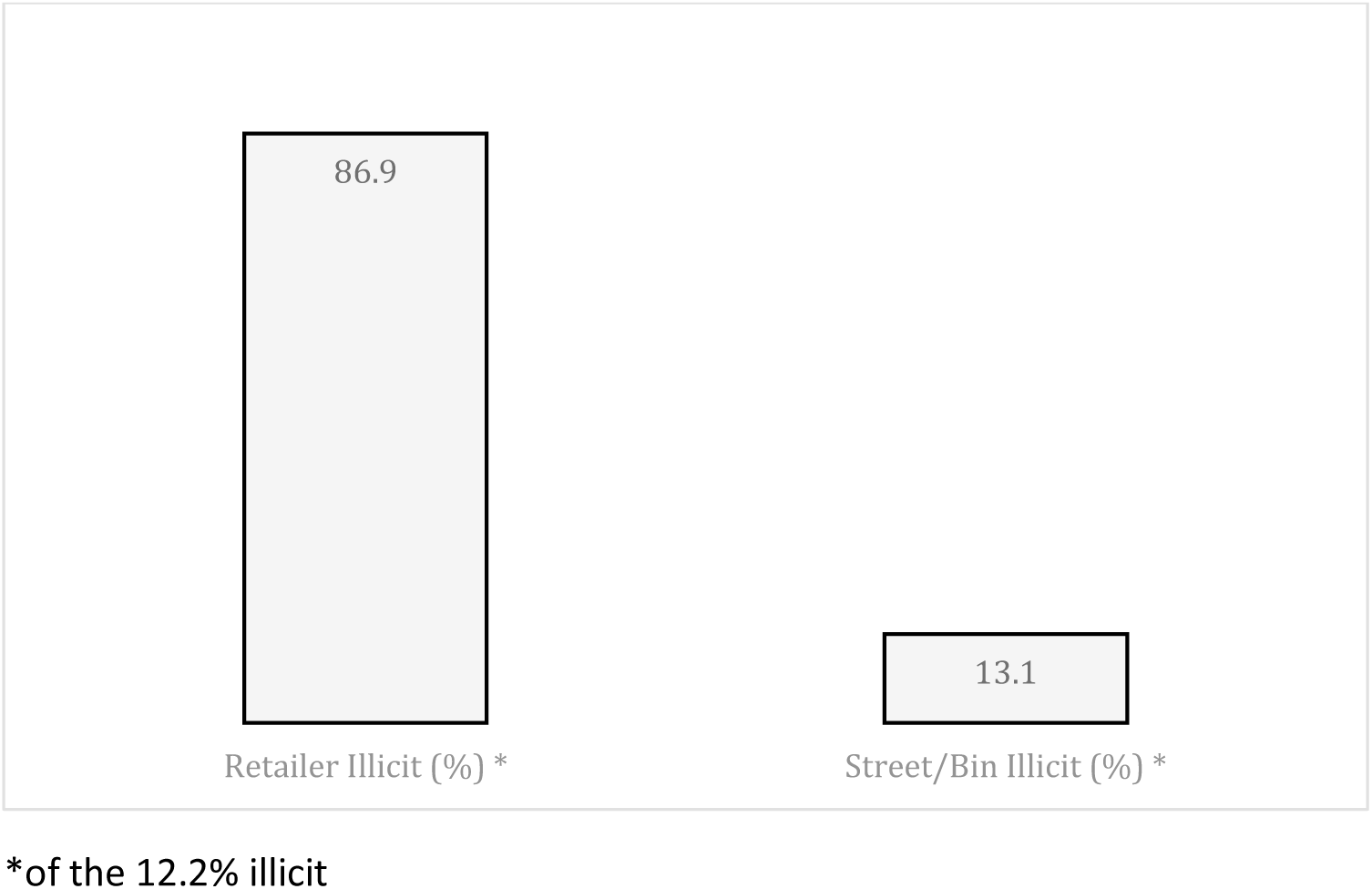
Distribution of the 12.2% illicit empty cigarette packs according to collection points

### Study Sites

The study was conducted in Mumbwa and Serenje (Central province), Chililabombwe, Kitwe, Mufulira, and Ndola (Copperbelt province), Chadiza, Chipata, and Katete (Eastern province), Mwense and Nchelenge (Luapula province), Luangwa, Lusaka, and Shibuyunji (Lusaka province), Isoka and Nakonde (Muchinga province), Chavuma and Mufumbwe (North-West province), Mbala and Nsama (Northern province), Chirundu, Livingstone, and Zimba (Southern province), and Mwandi and Sesheke districts (Western province) (Figure 1).

### Data Collection and Procedures

Data was collected by a team of 58 field staff including 25 supervisors and 33 research assistants (RAs), with two to six staff in each district. Supervisors were required to have a minimum qualification of a bachelor’s degree, while research assistants needed to have a minimum qualification of a General Certificate of Education (GCE) after completing grade 12. Field staff received three-day training consisting of: 1) classroom instruction on the data collection process, including recruitment of retailers, collection of empty cigarette packs from the retailers and streets/bins, and entering data in the questionnaire programmed on the tablets; and 2) field testing conducted in Chongwe district in Lusaka province. The field staff were deployed to their respective districts on the fourth day. Before any field work began, we made courtesy calls to key gatekeepers, including Provincial Administration Officers, Permanent Secretaries, District Commissioners, District Health Directors, Zambia Police Commands, Town Clerks/Council Chairpersons, Zambia Revenue Authority (ZRA) Senior Officials, and Market/Bus Terminus Chairpersons. The key gatekeepers responded overwhelmingly positively and pledged support within their jurisdictions to ensure the research team’s successful field implementation

The field work began with the mapping of main markets in each district and the listing of retailers in the area. The collection of empty cigarette packs was limited to retailers located within a 1km radius of the main market in the CBD of each district. RAs approached immobile retailers that were within a 1km radius of the CBD main market, i.e., the starting point of data collection for the district, and explained the aim of the study. Retailers who were interested in study participation were then provided with written study information and requested to sign a consent form if they agreed to participate. Retailers who provided written consent were supplied with a pre-labelled bag on the morning of pack collection day, and were asked to deposit all cigarette packs emptied throughout the day from single stick sales in the bag. The bags with the empty packs were retrieved in the evening of the same day. Each collection bag had a unique identifier and was color coded to distinguish between retailer and street/bin collections, with additional information on the geographical location. Along with providing the empty packs, the retailers also completed a short questionnaire about the cigarette brands they sold, their source, and buying (i.e., wholesale) and selling prices.

The collection of empty packs from the streets/bins began from the main market which was the starting point for data collection and covered all the streets/bins within the 1km radius. This was done for one day.

### Packs Processing

The RAs distributed pre-labelled bags to each retailer during their morning pack collection rounds and requested them to place all empty cigarette packs for the day inside. Bags with empty packs were retrieved in the evening. Each empty cigarette pack was placed in a plastic bag with a color code that distinguished it based on the source of collection (retailer or street/bin) with additional information of the geographical location. Each pack was examined for features such as brand name, flavor, cigarette size, pack size, local or imported, country of origin, manufacturer, the presence of textual health warning, compliance of the textual health warning in english, presence of ZRA tax stamp, and a duty-free stamp. These data were entered using the Open Data Kit (ODK) data collection tool and exported to Excel.

### Illicit Pack definition

A cigarette pack was considered illicit if it did not have any of the following features: 1) Textual health warning i.e., “TOBACCO IS HARMFUL TO HEALTH”; 2) textual health warning in English; and 3) a valid tax stamp from ZRA. Over and above the criteria above, a cigarette pack was also considered illicit if it had a duty-free stamp but was collected from a retailer who is not authorized to sell duty-free cigarettes. For packs collected from the streets or bins that displayed duty free stamps, we could not determine where the packs were purchased and therefore, we considered them legal for purposes of the study.

### Outcome and independent variables

#### Outcome variable

The outcome variables of interest was whether the cigarette pack was illicit or not. This was a binary outcome, where packs that met the illicit pack definition above were labelled “yes” for illicit cigarette pack, and those that did not were labelled “no”.

#### Independent variables

Regions (Rural vs Urban); Border status (border towns vs in-land towns); source type (imported or locally manufactured); Packing type (emperor packing, hinge-lid packing, shell slide packing; or soft packing), Economic zones (high income area vs low-income area), Variant flavor (full flavor, menthol or switch) Source (retailer vs street/bin) and the provinces.

### Data Analysis

We conducted descriptive statistics to estimate the overall proportion of illicit cigarettes (size of the illicit cigarette market), as well as for each province, district, and cigarette brand, and based on the source of the cigarette packs. Additionally, the proportion of illicit packs was calculated based on the specific criteria, i.e., whether the packs had a textual health warning, whether the warning was in English, and whether the pack had a duty-free stamp or a ZRA tax stamp. The frequencies were reported as numbers and percentages. The extent of tax evasion in Zambia was determined by focusing on the proportion of cigarette packs that did not have a ZRA tax stamp or had a duty-free stamp but was obtained from a retailer that is not authorized to sell duty-free cigarettes. Logistic regression was used to examine the factors associated with prevalence of illicit cigarette packs in unadjusted and adjusted models. Odds ratios (OR) and 95% confidence intervals (CI) were estimated from univariate and multivariable models. Despite other variables not reaching statistical significance, they were included a prior in the adjusted model as they were regarded important according to the literature. Statistical significance was set at the 0.05 level. The analyses were performed using STATA version 17.

## Results

### Descriptive Statistics

A total of 118, 344 empty cigarette packs were collected from the survey. Of these 81.9.0% (96,986) and 18.0% (21,358) were from retailers and streets/bins, respectively. Lusaka province had the highest collection 38.6% (45,652) seconded by Copperbelt province 18.9% (22,418) (supplementary 2). District wise, Lusaka district had the highest number of collected empty cigarette packs 35.9% (42,598), followed by Kitwe 6,31% (7469) and Nzama with 0.75% (883) with had the lowest collection (Table 1). Of the 3308 retail outlets, more than half 64% (2105) were kiosk/tumtembas and 15.6% (517) were brick stores (supplementary 3). In terms of brands, Rothmans accounted for 40.7% (48,172) packs of the total empty cigarette pack, Stuyvesant 13.1% (15,534) packs, wish 10.7% (12,641) packs, Pall Mall had 9.92% (11,734) packs (supplementary 4). The survey shows that the most expensive brand based on the retail price per pack was Dunhill (49.00 ZMW and 2.50 ZMW per stick), followed by Pall Mall (24.50 ZMW), Stuyvesant (23.60 ZMW), and Zark (22.42 ZMW) (supplementary table).

**Table 1:** Overall distribution of empty cigarette packs in Zambia by district and collection point (retailer and street/bin)

| Province & District | Retailer n (%) * | Street/Bin n (%) * | Total n (%) # |
| --- | --- | --- | --- |
| **Central** | **2987 (85.5)** | **505 (14.5)** | **3492 (2.95)** |
| Mumbwa | 2209 (90.7) | 226 (9.3) | 2435 (2.06) |
| Serenje | 778 (73.6) | 279 (26.4) | 1057 (0.89) |
| **Copperbelt** | **19379 (86.4)** | **3038 (13.6)** | **22418 (18.9)** |
| Chililabombwe | 3179 (87.7) | 445 (12.3) | 3625 (3.06) |
| Kitwe | 7164 (95.9) | 305 (4.1) | 7469(6.31) |
| Mufulira | 3347 (69.1) | 1498 (30.9) | 4845 (4.09) |
| Ndola | 5689 (87.8) | 790 (12.2) | 6479 (5.47) |
| **Eastern** | **10067 (74.4)** | **3459 (25.6)** | **13526 (11.4)** |
| Chadiza | 902 (70.2) | 382 (29.8) | 1284 (1.08) |
| Chipata | 7438 (79.7) | 1891 (29.3) | 9329 (7.90) |
| Katete | 1727 (59.3) | 1186 (40.7) | 2913 (2.46) |
| **Luapula** | **3567 (88.5)** | **462 (11.5)** | **4029 (3.40)** |
| Mwense | 2578 (98.0) | 52 (2.0) | 2630 (2.22) |
| Nchelenge | 989 (70.7) | 410 (29.3) | 1399 (1.18) |
| **Lusaka** | **39545 (86.6)** | **6107 (13.4)** | **45652 (38.6)** |
| Luangwa | 771 (43.4) | 1005 (56.6) | 1776 (1.50) |
| Lusaka | 37637 (88.4) | 4961 (11.6) | 42598 (35.9) |
| Shibuyunji | 1137 (89.0) | 141 (11.0) | 1278 (1.08) |
| **Muchinga** | **2990 (65.6)** | **1566 (34.4)** | **4556 (3.85)** |
| Isoka | 967 (79.5) | 250 (20.5) | 1217 (1.03) |
| Nakonde | 2023 (60.6) | 1316 (39.4) | 3339 (2.82) |
| **North-West** | **3321 (73.6)** | **1189 (26.4)** | **4510 (3.81)** |
| Chavuma | 1618 (63.5) | 932 (36.5) | 2550 (2.15) |
| Mufumbwe | 1703 (86.9) | 257 (13.1) | 1960 (1.66) |
| **Northern** | **2017 (76.3)** | **627 (23.7)** | **2644 (2.23)** |
| Mbala | 1481 (84.1) | 280 (15.9) | 1761 (1.49) |
| Nsama | 536 (60.7) | 347 (39.3) | 883 (0.75) |
| **Southern** | **10478 (77.1)** | **3104 (22.9)** | **13582 (11.5)** |
| Chirundu | 3276 (76.9) | 983 (23.1) | 4259 (3.60) |
| Livingstone | 4079 (80.2) | 1007 (19.8) | 5086 (4.30) |
| Zimba | 3123 (73.7) | 1114 (26.3) | 4237 (3.58) |
| **Western** | **2635 (66.9)** | **1301 (33.1)** | **3936 (3.32)** |
| Mwandi | 987 (69.5) | 434 (30.5) | 1421 (1.20) |
| Sesheke | 1648 (65.5) | 867 (34.5) | 2515 (2.13) |
| **Total** | **96986 (82.0)** | **21358 (18.0)** | **118344** |

Footnote: \* Row %; #Column %

### Description of illicit empty packs collected

Of the 118,344 cigarette packs collected, 12.2% (14,428) were illicit (Figure 1). Western province had the highest proportion of illicit cigarette packs, and of the 3,936 empty packs collected 32% (1,260) were illicit (Table 2). Lusaka province had the second highest proportion of illicit cigarette packs. Out of the 45,652 packs collected in the province, 24.6% (11,233) packs were illicit. When classified under inland and border towns, 31.8% (37,671) were collected from the nine border districts and 5.0% (1,897) were classified as illicit. On the other hand, 15.5% (12,531) of the total 80,673 packs collected in the non-border districts were classified as illicit. When we stratified into urban and rural, 70,961 packs collected in the urban, and of these 15.3% (10.876) were classified as illicit (Table 2). In contrast, the rural districts accounted for 7.5% (3,552) illicit packs out of the total of 47,383 packs collected from those districts.

**Table 2:** Distribution of licit and illicit cigarette packs by province and district.

| Province/District | Retailer legal (%) * | Retailer Illicit (%) * | Street/Bin legal (%) * | Street/Bin Illicit (%) * | All Legal (%) * | All Illicit (%) * | Total Packs |
| --- | --- | --- | --- | --- | --- | --- | --- |
| **Central** | **2881 (96.5)** | **106 (3.5)** | **491 (97.2)** | **14 (2.8)** | **3372 (96.6)** | **120 (3.4)** | **3492** |
| Mumbwa | 2123 (96.1) | 86 (3.9) | 212 (93.8) | 14 (6.2) | 2335 (95.9) | 100 (4.1) | 2435 |
| Serenje | 758 (97.4) | 20 (2.6) | 279 (100.0) | 0 (0.0) | 1037 (98.1) | 20 (1.9) | 1057 |
| **Copperbelt** | **19291 (99.5)** | **88 (0.5)** | **3034 (99.9)** | **4 (0.1)** | **22325 (99.6)** | **92 (0.4)** | **22417** |
| Chililabombwe<sup>&</sup> | 3155 (99.2) | 24 (0.8) | 444 (99.8) | 1 (0.2) | 3599 (99.3) | 25 (0.7) | 3624 |
| Kitwe<sup>+</sup> | 7118 (99.4) | 46 (0.6) | 302 (99.0) | 3 (1.0) | 7420 (99.3) | 49 (0.7) | 7469 |
| Mufulira<sup>&</sup> | 3347 (100.0) | 0 (0.0) | 1498 (100.0) | 0 (0.0) | 4845 (100) | 0 (0.0) | 4845 |
| Ndola<sup>+</sup> | 5671 (99.7) | 18 (0.3) | 790 (100.0) | 0 (0.0) | 6461 (99.7) | 18 (0.3) | 6479 |
| **Eastern** | **9943 (98.8)** | **124 (1.2)** | **3459 (100.0)** | **0 (0.0)** | **13402 (99.1)** | **124 (0.9)** | **13526** |
| Chadiza | 862 (95.6) | 40 (4.4) | 382 (100.0) | 0 (0.0) | 1244 (96.9) | 40 (3.1) | 1284 |
| Chipata<sup>&+</sup> | 7376 (99.2) | 62 (0.8) | 1891 (100.0) | 0 (0.0) | 9267 (99.3) | 62 (0.7) | 9329 |
| Katete<sup>&</sup> | 1705 (98.7) | 22 (1.3) | 1186 (100.0) | 0 (0.0) | 2891 (99.2) | 22 (0.8) | 2913 |
| **Luapula** | **2845 (79.8)** | **722 (20.2)** | **460 (99.6)** | **2 (0.4)** | **3305 (82.0)** | **724 (18.0)** | **4029** |
| Mwense | 1869 (72.5) | 709 (27.5) | 52 (100.0) | 0 (0.0) | 1921 (73.0) | 709 (27.0) | 2630 |
| Nchelenge | 976 (98.7) | 13 (1.3) | 408 (99.5) | 2 (0.5) | 1384 (98.9) | 15 (1.1) | 1399 |
| **Lusaka** | **29393 (74.3)** | **10152 (25.7)** | **5026 (82.3)** | **1081 (17.7)** | **34419 (75.4)** | **11233 (24.6)** | **45652** |
| Luangwa | 758 (98.3) | 13 (1.7) | 994 (98.9) | 11 (1.1) | 1752 (98.6) | 24 (1.4) | 1776 |
| Lusaka<sup>+</sup> | 27944 (74.2) | 9693 (25.8) | 3961 (79.8) | 1000 (20.2) | 31905 (74.9) | 10693 (25.1) | 42598 |
| Shibuyunji | 691 (60.8) | 446 (39.2) | 71 (50.4) | 70 (49.6) | 762 (59.6) | 516 (40.4) | 1278 |
| **Muchinga** | **2964 (99.1)** | **26 (0.9)** | **1559 (99.6)** | **7 (0.4)** | **4523 (99.3)** | **33 (0.7)** | **4556** |
| Isoka | 941 (97.3) | 26 (2.7) | 250 (100.0) | 0 (0.0) | 1191 (97.9) | 26 (2.1) | 1217 |
| Nakonde<sup>&</sup> | 2023 (100.0) | 0 (0.0) | 1309 (99.5) | 7 (0.5) | 3332 (99.8) | 7 (0.2) | 3339 |
| **North-West** | **3280 (98.8)** | **41 (1.2)** | **1188 (99.9)** | **1 (0.1)** | **4468 (99.1)** | **42 (0.9)** | **4510** |
| Chavuma | 1613 (99.7) | 5 (0.3) | 932 (100.0) | 0 (0.0) | 2545 (99.8) | 5 (0.2) | 2550 |
| Mufumbwe | 1667 (97.9) | 36 (2.1) | 256 (99.6) | 1 (0.4) | 1923 (98.1) | 37 (1.9) | 1960 |
| **Northern** | **1978 (98.1)** | **39 (1.9)** | **626 (99.8)** | **1 (0.2)** | **2604 (98.5)** | **40 (1.5)** | **2644** |
| Mbala<sup>&</sup> | 1468 (99.1) | 13 (0.9) | 280 (100.0) | 0 (0.0) | 1748 (99.3) | 13 (0.7) | 1761 |
| Nsama | 510 (95.1) | 26 (4.9) | 346 (99.7) | 1 (0.3) | 856 (97.0) | 27 (3.0) | 883 |
| **Southern** | **9942 (94.9)** | **536 (5.1)** | **2880 (92.8)** | **224 (7.2)** | **12822 (94.4)** | **760 (5.6)** | **13582** |
| Chirundu<sup>&</sup> | 2793 (85.3) | 483 (14.7) | 773 (78.6) | 210 (21.4) | 3566 (83.7) | 693 (16.3) | 4259 |
| Livingstone<sup>&+</sup> | 4038 (99.0) | 41 (1.0) | 994 (98.7) | 13 (1.3) | 5032 (99.0) | 54 (1.0) | 5086 |
| Zimba | 3111 (99.6) | 12 (0.4) | 1113 (99.9) | 1 (0.1) | 4224 (99.7) | 13 (0.3) | 4237 |

| **Province/District** | **Retailer legal (%) \*** | **Retailer Illicit (%) \*** | **Street/Bin legal (%) \*** | **Street/Bin Illicit (%) \*** | **All Legal (%) \*** | **All Illicit (%) \*** | **Total Packs** |
| --- | --- | --- | --- | --- | --- | --- | --- |
| **Western** | **1915 (72.7)** | **720 (27.3)** | **761 (58.5)** | **540 (41.5)** | **2676 (68.0)** | **1260 (32.0)** | **3936** |
| Mwandi | 816 (82.7) | 171 (17.3) | 366 (84.3) | 68 (15.7) | 1182 (83.0) | 239 (17.0) | 1421 |
| Sesheke<sup>&</sup> | 1099 (66.7) | 549 (33.3) | 395 (45.6) | 472 (54.4) | 1494 (59.0) | 1021 (41.0) | 2515 |
| **Total** | **84432 (71.3)** | **12554 (10.6)** | **19484 (16.5)** | **1874 (1.6)** | **103916 (87.8)** | **14428 (12.2)** | **118344** |

Footnote: \*Row %; <sup>&</sup>border towns

### Textual warning, Duty-free stamp and ZRA tax stamp

The empty packs examination showed that 1.5% (1792) had no textual health warning, 0.3 (343) had textual health warning in another language apart from English. 1.5% (1490) had a duty-free stamp, and 10.1% (11939) had no ZRA stamp (Table 3)

**Table 3:**
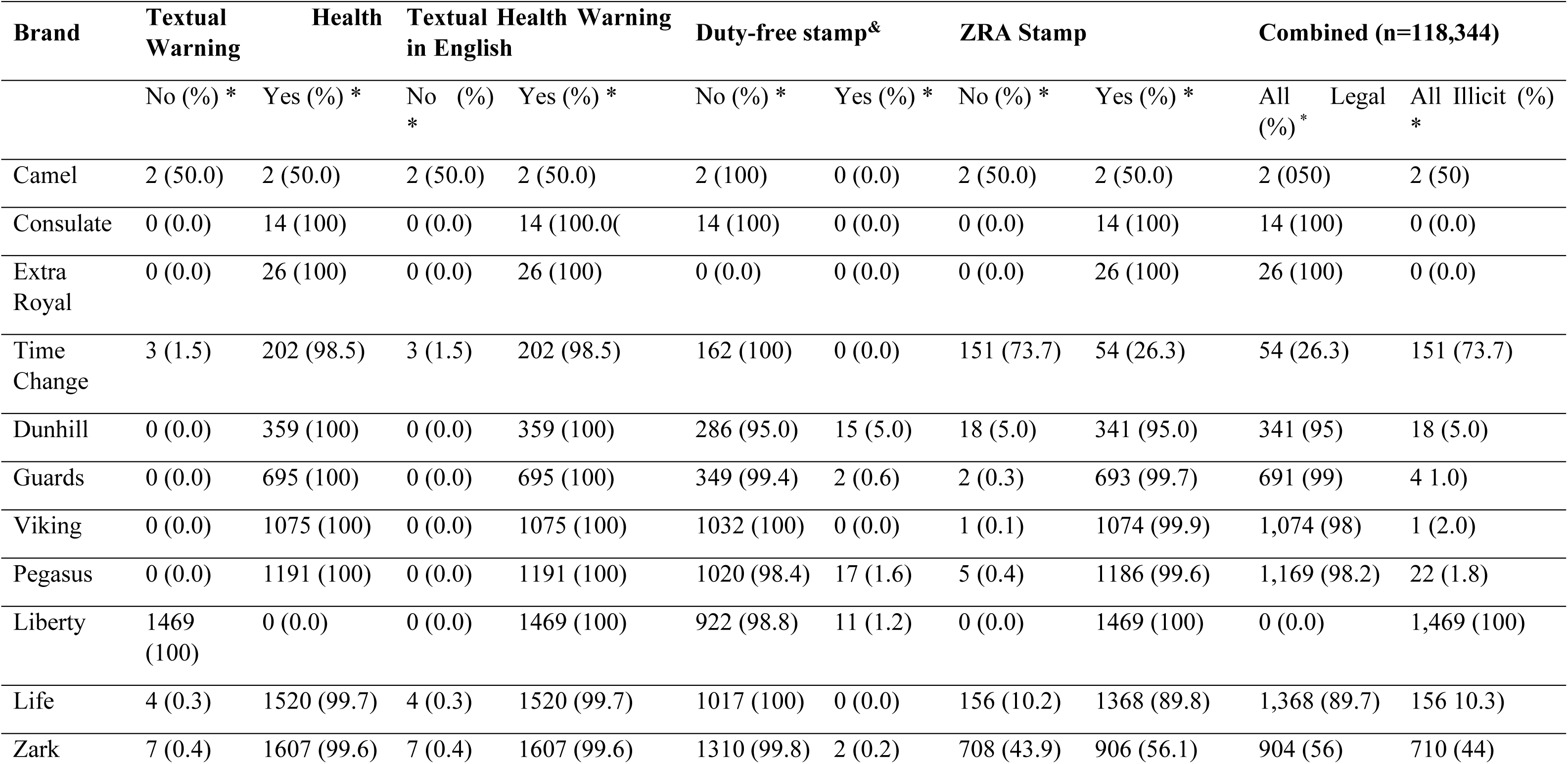
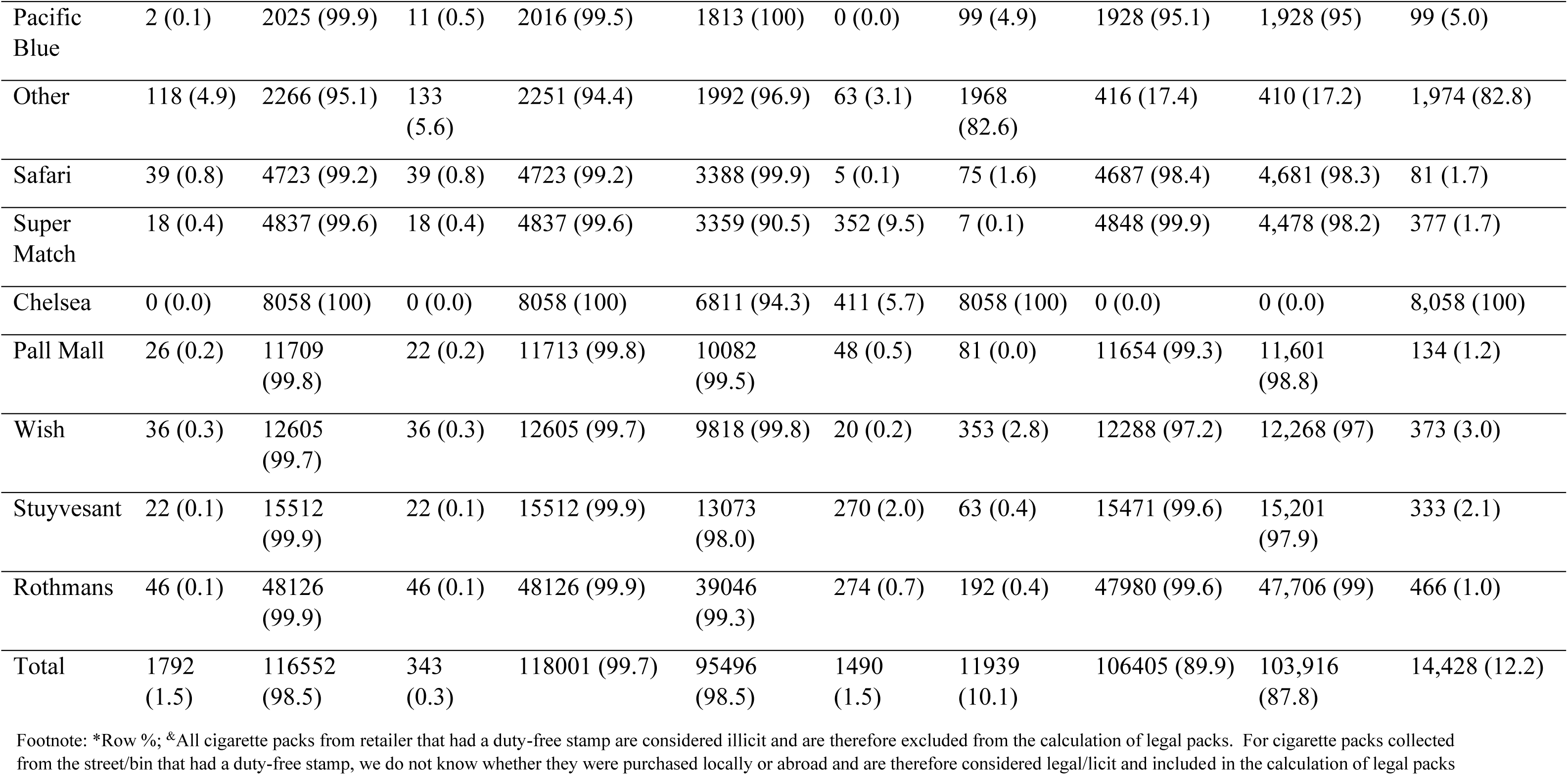
Compliance of cigarette brands based on criterion.

### Factors associated with illicit cigarette packs

Table 4, shows the results from the unadjusted and adjusted logistic regression model of factors associated with illicit cigarettes packs in Zambia.

**Table 4:** Unadjusted and adjusted logistic regression model for factors associated with illicit cigarette in Zambia.

| **Characteristic** | **UOR\*** | **95% CI** | **AOR\*\*** | **95% CI** |
| --- | --- | --- | --- | --- |
| **Region** |  |  |  |  |
| Rural | Ref | Ref | Ref | Ref |
| Urban | 2.53 | 2.43 – 2.65 | 2.23 | 2.17 – 2.43 |
| **Border status** |  |  |  |  |
| Border towns | Ref | Ref | Ref | Ref |
| In-land towns | 3.26 | 3.11 – 3.44 | **0.17** | **0.13 – 0.23** |
| **Source type** |  |  |  |  |
| Imported | Ref | Ref | Ref | Ref |
| Local | 0.39 | 0.36 – 0.45 | **0.44** | **0.37 – 0.53** |
| **Packing type** |  |  |  |  |
| Emperor Pack | Ref | Ref | Ref | Ref |
| Hinge-lid | 0.39 | 0.36 – 0.43 | **0.55** | **0.46 – 0.65** |
| Shell slide pack | 0.008 | 0.005 – 0.01 | **0.04** | **0.02 – 0.07** |
| Soft pack | 0.03 | 0.02 – 0.09 | 1.38 | **1.11 – 1.71** |
| **Outlet type** |  |  |  |  |
| Cigarette stand | Ref | Ref | Ref | Ref |
| General store | 1.86 | 1.72 – 2.01 | 11.7 | **9.88 – 13.8** |
| Kiosk | 1.47 | 1.37 – 1.59 | 6.44 | **5.64 – 7.35** |
| **Economic management** |  |  |  |  |
| High income area | Ref | Ref | Ref | Ref |
| Low-income area | 0.77 | 0.74 – 0.80 | 1.11 | 0.99 – 1.24 |
| **Variant flavor** |  |  |  |  |
| Full flavor | Ref | Ref | Ref | Ref |
| Menthol | 3.97 | 3.81 – 4.12 | 1.21 | **1.06 – 1.35** |
| Switch | 12.91 | 11.43 – 14.71 | 1.72 | **1.34 – 2.21** |
| **Source** |  |  |  |  |
| Retailer | Ref | Ref | Ref | Ref |
| Street/bin | 0.71 | 0.67 – 0.75 | **0.31** | **0.25 – 0.38** |
| **Province** |  |  |  |  |
| Central | Ref | Ref | Ref | Ref |
| Copperbelt | 0.11 | 0.08 – 0.15 | **0.17** | **0.10 – 0.27** |
| Eastern | 0.21 | 0.15 – 0.29 | **0.22** | **0.15 – 0.36** |
| Luapula | 0.05 | 0.02 – 0.12 | **0.12** | **0.04 – 0.38** |
| Lusaka | 10.2 | 8.23 – 12.5 | **4.11** | **2.91 – 5.81** |
| Muchinga | 0.06 | 0.03 – 0.13 | **0.03** | **0.004 – 0.02** |
| North-Western | 0.35 | 0.24 – 0.51 | **0.52** | **0.32 – 0.81** |
| Northern | 0.54 | 0.37 – 0.79 | 1.48 | 0.93 – 2.37 |
| Southern | 2.10 | 1.63 – 2.63 | 1.16 | 0.77 – 1.74 |
| Western | 17.1 | 13.7 – 21.3 | **0.35** | **0.23 – 0.53** |

UOR\* Unadjusted odds ratio; AOR\*\* Adjusted odds ratio

Location, packaging, the point-of-sale cigarettes variables significantly explain the prevalence of the illicit cigarette market. Empty cigarette packs collected from the urban regions had 2.25 higher odds of being illicit than the cigarette packs collected from the rural regions. Except other provinces, Lusaka, Northern and Southern provinces had higher odds of having illicit cigarette packs than the central province. The cigarette packs from the in-land towns had 0.17 lower odds of being illict than those collected from the towns near the border. In terms of sources, the local cigarettes had 0.44 lower odds of being illicit than the imported cigarettes, implying the imported cigarettes were more likely to be illicit than the domestic ones.

Packaging wise, cigarette pack packaged Hinge-lid and Shell slide have lower odds of being illicit than the emperor packaging, while cigarettes in soft pack have higher odds of being illicit than the emperor packaging. Cigarettes that were sold in general stores and kiosks were had 11.7 and 6.44, respectively higher odds of being illicit than those sold in cigarette stands. Cigarette collection from the street/bins had 0.31 lower odds of being illicit than those collected from the retailers. Cigarettes that were in menthol and switch flavors had higher odds of being illicit than those that were in full flavors.

## Discussion

To the best of our knowledge, this study is the first to estimate the proportion of cigarettes consumed that are illicit, the extent of cigarette tax evasion, and the associated factors in Zambia. We estimated that 12.2% of cigarettes consumed in Zambia are illicit. One notable contributor to illicit cigarette trade in Zambia was tax evasion: 10.9% of the collected packs did not have the ZRA tax stamp suggesting that they had not paid the required tax, and/or had a duty-free stamp but were collected from retailers that were not authorized to sell duty-free cigarettes. Non-compliance with textual health warning requirements was low: only 1.55% either did not have a textual health warning or had one that was not in English. We also found that packs that were collected from inland districts, streets/bins, and those that were for locally manufactured cigarettes were less likely to be illicit than those collected from border districts, retailers and those that were for imported cigarettes, respectively. On the other hand, packs from urban areas, and general stores or kiosks were more likely to be illicit than those collected from rural areas, and those from cigarette stands, respectively.

Our results are comparable to those from studies that are independent of the tobacco industry conducted in a number of other African countries which have reported prevalence of illicit trade cigarettes ranging from approximately 9% (in the Gambia)^15^ to 20% (in Ghana)^16^. Our results are also consistent with global estimates on average the illicit cigarette trade market is about 16.8% for low-income countries^17^. Studies in Ethiopia and South Africa, on the other hand, reported a prevalence of illicit cigarettes of 45.4%^18^ and 54%^19^, respectively and these are much higher than what we found in Zambia. We found that the prevalence of illicit cigarette packs was much higher among imported packs at (61.7%) when compared to locally manufactured cigarettes (3.2%). In addition, in line with other studies ^16,18^, we found large regional differences in the prevalence of illicit cigarettes, from 0.4% in Copperbelt to 32% in the Western province, and that border areas are more vulnerable to illicit cigarettes than inland areas. Considering that taxes are the same throughout Zambia, our findings support the argument that illicit cigarettes are not linked to the level of taxation or higher prices, but weak control systems at the borders which allows smuggling of illicit cigarettes from other countries^16^.

The brands with the highest proportion of illicit cigarette packs were Chelsea (100%), Liberty (100%) and Time Change (74%). Although non-compliance with textual health warning requirements was low, this was 100% for Liberty, a locally manufactured brand. Most of the packs that did not comply with textual health warnings had a ZRA tax stamp, which might indicate the need to strengthen collaboration and coordination between ZRA and the relevant health departments responsible for textual health warning inspections. On the other hand, 100% of the Chelsea packs, a brand originating from Zimbabwe and not registered for importation by the ZRA, were non-compliant with the ZRA tax stamp requirements. This validates that Zimbabwe which is considered the primary source of illicit cigarettes in Southern Africa, is also the source of cigarette smuggling in Zambia^20,21^.

Illicit cigarettes have been found to be generally cheaper than legal cigarettes. If the illicit cigarette market is left unchecked, this might result in increases in the smoking prevalence and revenue losses for the government. Therefore, it is important to take the appropriate actions to control the illicit cigarette market and to monitor it regularly. To counter the supply of illicit cigarettes, Zambia should urgently secure its cigarette supply chain, including the introduction of a secure track and trace system for cigarettes. Kenya, for example, managed to reduce its illicit cigarette market from 15% in 2003 to 5% in 2016 through a comprehensive strategy that included sticking tax stamps on cigarettes for domestic consumption, implementing a track-and-trace system and introducing scanners and point of entry into the country^22^. For Zambia, this could also include building capacity by training enough immigration, police and customs officers and intensifying the patrols along the borders. Zambia is also yet to ratify the WHO Protocol on Illicit Tobacco Trade. The ratification of the WHO Protocol on Illicit Tobacco Trade would indicate a political commitment to a systematic effort to combat the illicit trade within Zambia.

Our study was dependent on the retailers providing empty packs and it is possible that they might have kept some illicit packs due to fear of prosecution. This would have resulted in underestimation of the proportion of illicit cigarette packs. In addition, any packs that were collected from the street that had a duty-free stamp could not be classified as illicit. However, this was a very small proportion of packs and would not have made a big difference to the results and their interpretation. Despite adjusting for known confounders in the multivariable model, the potential for the residual confounders inherent in observational studies remains and might affect the interpretation of study outcomes One of the major strengths of this study is the large sample size and the ability to generalize the results at a country level. The data was collected in a nationally representative manner, encompassing both border and non-border districts, urban and rural areas, different market conditions, and including the largest cities in Zambia. Additionally, this study provides the first contribution to understanding illicit cigarette trade in Zambia and identifies the key brands most notorious for such trade. It also highlights the locations where these illicit cigarettes are predominantly sold and emphasizes that tax evasion is the primary contributor to this trade.

## Conclusion

Although the prevalence of illicit cigarettes is low in Zambia at 12.2%, it is still important for the country to secure its cigarette supply chain. Western provinces had the highest proportion of illicit cigarette packs with most of them being imported. These findings underscore the fact that Zambia should ratify and implement the WHO Protocol on Illicit Tobacco Trade to counter the supply of illicit cigarettes. Adoption of a track and trace system would enable customs officers to detect counterfeit and smuggled cigarettes. Furthermore, there is a need to increase capacity building in training enough immigration, police and customs officers and intensify the patrols along the borders.

## Ethics Statements

Patient consent for publication Not applicable

## Ethics approval

Ethics approval was obtained from the University of Zambia Biomedical Research Ethics Committee (UNZABREC) (REF 3288-2022) and permission to conduct the study was obtained from the National Health Research Authority (NHRA) (REF-NHRA 0000002/2/11/2022). Courtesy calls and introductory letters were written to ZRA offices, District Commissioners of each district, and the Zambia Police district office.

## Data Availability

All data produced in the present work are contained in the manuscript

The data sets used and/ or analyzed during the current study is available at the zenodo link **10.5281/zenodo.10993175**

## Acknowledgements

This research was carried out within the framework of the Tobacco Control Data Initiative (TCDI). TCDI is a project that encompasses six African nations (the Democratic Republic of Congo, Ethiopia, Kenya, Nigeria, South Africa, and Zambia). Its objective is to comprehend the tobacco-related data requirements of these countries, assess existing data, pinpoint gaps in available tobacco data, gather new data to address those gaps, and create tools to assist policymakers in utilizing crucial data more effectively for shaping tobacco policies. The initiative is spearheaded by Development Gateway: an IREX Venture (DG), a global nonprofit organization specializing in data-driven development, in collaboration with the University of Cape Town’s Research Unit on the Economics of Excisable Products (REEP). We extend our appreciation to Center for Primary Care Research based in Lusaka, Zambia for leading the data collection and analysis on behalf of DG. We are also grateful for the support extended by key stakeholders in Zambia’s tobacco control sector. In particular, we would like to acknowledge Provincial Administration Officers, Permanent Secretaries, District Commissioners, District Health Directors, Zambia Police Commands, Town Clerks/Council Chairpersons, Zambia Revenue Authority (ZRA) Senior Officials, and Market/Bus Terminus Chairpersons and the Ministry of Health.

## Contributors

**CZ:** Conceptualization, Methodology, Zonal lead data collection, Analyzed the data, interpreted the findings, writing – original draft, writing – review & editing. **MP:** Conceptualization, Methodology, Zonal lead data collection, Analyzed the data, interpreted the findings, review & editing. **WM:** Conceptualization, Methodology, Zonal lead data collection, review & editing. **RZ:** Conceptualization, Methodology, Zonal lead data collection, interpreted the findings, review & editing. **MM:** Conceptualization, Methodology, data analysis, review & editing. **AC:** conceptualization, review & editing. **SO**: conceptualization, data analysis, review & editing. **SA:** conceptualization, review & editing, **RP**: data analysis, review & editing. **HR:** Conceptualization, Methodology, interpreted the findings, review & editing. **FG:** Conceptualization, Methodology, interpreted the findings, review & editing. **NM:** Conceptualization, Methodology, Analyzed the data, interpreted the findings, review & editing, and project oversight. All authors approved the final version of the manuscript.

## Funding

This study received support from the Bill & Melinda Gates Foundation (INV-009670). However, the funders were not involved in designing the study, gathering or analyzing data, deciding to publish, or preparing the manuscript. The opinions and conclusions presented here are solely those of the authors and do not necessarily represent the views or policies of the Bill & Melinda Gates Foundation.

## Competing interests

The authors have declared that no competing interests exist

## Patient and public involvement

We actively engaged key tobacco control actors in Zambia in developing the research question(s) and ensuring these were informed by their priorities, experience, and preferences. These key actors were also involved in the design of the study and interpretation of the results. In November and December 2021, we held one-on-one in-person interviews with 18 individuals (five government officials, ten civil society organizations, two academics and one development partner), who represented 15 organizations that are key tobacco control actors and stakeholders in Zambia. These discussions aimed to understand their tobacco control data needs, identify existing data, confirm gaps in the available data, and what they perceived as the priority gaps for immediate research. Research on illicit trade of cigarettes in Zambia emerged as a research priority from this process. We then convened an in-person validation workshop in February 2022 whose objectives included confirming the key actors’ choice on the research priority and refining the specific questions that needed to be addressed, and the study design and methods to be used. In July 2023, after completion of data collection and analysis, we held an online workshop where we presented our findings to the tobacco control actors in Zambia, and obtain feedback on the way that we had interpreted them.

## Provenance and peer review

Not commissioned; externally peer reviewed

## Notes

### Competing Interest Statement

The authors have declared no competing interest.

### Funding Statement

Melinda and Bill Gates

### Author Declarations

Ethics approval was obtained from the University of Zambia Biomedical Research Ethics Committee (UNZABREC) (REF 3288-2022) and permission to conduct the study was obtained from the National Health Research Authority (NHRA) (REF-NHRA 0000002/2/11/2022).

## Reference

1. Mathers CD, Loncar D. Projections of global mortality and burden of disease from 2002 to 2030. PLoS Med. Nov 2006;3(11):e442.

2. Sumithrarachchi SR JR, Warnakulasuriya S.. Betel Quid Addiction: A Review of Its Addiction Mechanisms and Pharmacological Management as an Emerging Modality for Habit Cessation. Subst Use Misuse. 2021;56(13):2017–2025.

3. WHO. Zambia Steps Survey for Non Communicable Diseases. Zambia Report for 2017 Zambia Steps Survey for Non Communicable Diseases. Zambia Report for 2017. WHO. 2017.

4. Olowski P MC. Differential burden and determinants of tobacco smoking: population-based observations from the Zambia demographic and health survey (2002 and 2007). J Healthc Commun. 2015;1:2472–1654.

5. Cosmas Zyambo OB, Peter Songolo, Adamson S Muula, Emmanuel Rudatsikira, Seter Siziya. Prevalence and predictors of smoking in a mining town in Kitwe, Zambia: A 2011 population-based survey. Health. 2013;5(6):1021–1025.

6. WHO. Tobacco control as an accelerator for the Sustainable Development Goals https://fctc.who.int/publications/m/item/tobacco-control-accelerator-sustainable-development-goals-zambia (accessed 08_04_24). *WHO FCTC.* 2021.

7. T T. Zambia-Country Profile, Tobacco Tactics, updated 13 September 2021 (accessed 8_04_24). Bath University 2021.

8. Guindon GE, Tobin S, Yach D. Trends and affordability of cigarette prices: ample room for tax increases and related health gains. Tob Control. Mar 2002;11(1):35–43.

9. Felsinger R, Groman E. Price Policy and Taxation as Effective Strategies for Tobacco Control. Front Public Health. 2022;10:851740.

10. Paraje G, Stoklosa M, Blecher E. Illicit trade in tobacco products: recent trends and coming challenges. Tob Control. Mar 2022;31(2):257–262.

11. Bader P, Boisclair D, Ferrence R. Effects of tobacco taxation and pricing on smoking behavior in high risk populations: a knowledge synthesis. Int J Environ Res Public Health. Nov 2011;8(11):4118–4139.

12. WHO. Protocol to Eliminate Illicit Trade in Tobacco Products, https://fctc.who.int/protocol/overview (accessed 8_04_24). *WHO.* 2013.

13. B Z. Roland Imperial Tobacco Company Calls for Ban on Importation of Cigarettes, https://web.facebook.com/RadioPhoenixZambia/photos/a.134442918747/10160038281578748/?type=3&_rdc=1&_rdr (accessed 08_04_24). Phoenix FM Zambia. 2022.

14. N C. 400 million cigarettes enter Zambia illegally, https://zambianbusinesstimes.com/400-million-cigarettes-enter-zambia-illegally/ *Zambian Business Times.* 2020.

15. Chisha Z, Janneh ML, Ross H. Consumption of legal and illegal cigarettes in the Gambia. Tob Control. Oct 2020;29(Suppl 4):s254–s259.

16. Singh A, Ross H, Dobbie F, et al. Extent of illicit cigarette market from single stick sales in Ghana: findings from a cross-sectional survey. BMJ Open. Mar 22 2023;13(3):e062476.

17. Joossens L, Merriman D, Ross H, Raw M. The impact of eliminating the global illicit cigarette trade on health and revenue. Addiction. Sep 2010;105(9):1640–1649.

18. Dauchy E RH. Is Illicit Cigarette Market a Threat to Tobacco Control in Ethiopia?. Nicotine Tob Res. 2022;24(8):1228–1233.

19. Vellios N, van Walbeek C, Ross H. Illicit cigarette trade in South Africa: 2002-2017. Tob Control. Oct 2020;29(Suppl 4):s234–s242.

20. Haysom S. The Illicit Tobacco Trade in Zimbabwe and South Africa: Impacts and Solutions. Atlantic Council: Scowcroft Centre for Strategy and Security Working Papers. 2019.

21. Shaw M, & Reitano, T. Case Studies of Illicit Trade In Developing Economies. Atlantic Council. https://www.jstor.org/stable/pdf/resrep29474.7.pdf *(11_04_24)*. 2020.

22. Ross H. Tracking and tracing tobacco products in Kenya. Prev Med. Dec 2017;105S:S15–S18.

